# Methods for safely sharing dual-use genetic data

**DOI:** 10.1101/2024.11.29.24318203

**Authors:** Sterling Sawaya, Chien-Chi Lo, Po-E Li, Blake Hovde, Patrick Chain

## Abstract

**Abstract:** *Background:* Some genetic data has dual-use potential. Sharing pathogen data has shown tremendous value. For example therapeutic development and lineage tracking during the COVID pandemic. This data sharing is complicated by the fact that these data have the potential to be used for harm. The genome sequence of a pathogen can be used to enable malicious genetic engineering approaches or to recreate the pathogen from synthetic DNA. Standard data security methods can be applied to genetic data, but when data is shared between institutions, ensuring appropriate security can be difficult. Sensitive data that is shared internationally among a wide array of institutions can be especially difficult to control. Methods for securely storing and sharing genetic data with potential for dual-use are needed to mitigate this potential harm.

*Results:* Here we propose new methods that allow genetic data to be shared in a data format that prevents a nefarious actor from accessing sensitive aspects of the data. Our methods obfuscate raw sequence data by pooling reads from different samples. This approach can ensure that data is secure while stored and during electronic transfer. We demonstrate that by pooling raw sequence data from multiple samples of the same organism, the ability to fully reconstruct any individual sample is prevented. In the pooled data, most genomic information remains, but reads or mutations cannot be directly attributed to any individual sample. To further restrict access to information, regions of a genome can be removed from the reads.

*Conclusion:* Our methods obscure genomic information within raw sequence reads. This method can allow genetic data to be stored and shared while preventing a nefarious actor from being able to perfectly reconstruct an organism. Broad-scale sequence information remains, while fine scale details about specific samples are difficult or impossible to reconstruct.

## Background

As biotechnology advances, there is an increased potential that it can be misused [1]. Today, methods in synthetic biology can facilitate the creation of novel organisms. DNA can be accurately synthesized and this DNA can be inserted into an organism using CRISPR/CAS9 [2]. Entire portions of a genome can either be directly synthesized or be rewritten to become any desired sequence. Furthermore, entire organisms can be constructed with synthetic DNA [3]. The ability to fully reconstruct a virus or bacteria with only a genome sequence has been demonstrated [4]. Although today this method is more cumbersome and expensive than simply adding or deleting genes from a genome, as synthetic biology continues to advance, these capabilities will become easier and more widespread.

The ability to recreate an organism with genetic data introduces a dilemma for those generating pathogen genome data. There is a need to share genetic information of pathogens so that their origin and evolution can be understood, and also a beneficial use of the genetic information when designing countermeasures to an infectious disease [5,6]. However, because this data can be misused, there may be hesitance to share genetic data that has been collected [7], or perhaps even withhold publishing to avoid pressure to share the data. For genetic data from the most dangerous pathogens, sharing data may be prohibited in some countries, especially if the data is shared internationally [8,9].

There are international treaties for sharing specific pathogen data, such as the Pandemic Influenza Preparedness Framework [10]. This framework encourages the sharing of genomic data from influenza, but is, however, not legally binding and its enforceability has never been tested [11]. There are ongoing negotiations to expand this approach to other pathogen data, but in general the sharing of pathogen sequence data only occurs between countries through bilateral agreements [12]. The ownership of genetic data and the sharing of benefits or products from genetic data are governed by the Nagoya protocol, but this treaty requires a case-by-case bilateral negotiation to make specific arrangements [13].

The standards and regulations for data sharing during a world-wide crisis have been tested by the COVID pandemic. During the initial stages of the outbreak, sharing of pathogen sequence data was essential for tracking its spread, predicting how the virus may evolve, and developing countermeasures such as mRNA vaccines [14,15]. Without the prompt sharing of accurate sequence data, the world may not have been able to respond rapidly to this novel disease. Nevertheless, as data was openly shared, concerns about equity and benefit-sharing arose [16,17]. These and other concerns can restrict fully open pathogen genomic data sharing during disease outbreaks.

The open sharing of scientific data stands as an ideal standard on which major scientific advances rely [18,19]. However, this data sharing approach can conflict with the data sharing restrictions in place for the genomic data of certain pathogens. Currently, there is no standard method by which critical data from dangerous pathogens can be shared that is commensurate with requisite security. The data is either siloed, so no genetic data is shared, or the data is wholly shared, allowing anyone with access to the data to recreate the pathogen [20]. New, safer methods for accessing the sequence data of pathogens are needed.

The methods here are inspired by techniques in molecular cryptography, in which cryptographic protocols are applied directly to DNA molecules to ensure that sensitive information is protected before the DNA is sequenced. Molecular cryptography can be used to mix DNA samples in such a way that a key is needed to determine which read from a DNA sequencer belongs to which sample [21]. Without the key, data from the resulting pool is obfuscated. A similar approach can be done with software, in which reads from multiple files are pooled together and the meta-data is stripped. Using software, we examine the use of this approach. Here, we propose a new method that allows for sharing genetic sequence data while protecting sensitive aspects of the data.

## Methods

We developed new methods for formatting genetic data that prevents anyone with access to the data from being able to reconstruct an entire genome (Figure 1). The first proposed method involves the following (Figure 1B): DNA sequencing reads from a sample (or group of samples) are aligned to an incomplete reference genome. A substantial portion of this genome must be removed (e.g. removal of a full gene).

**Figure 1.**
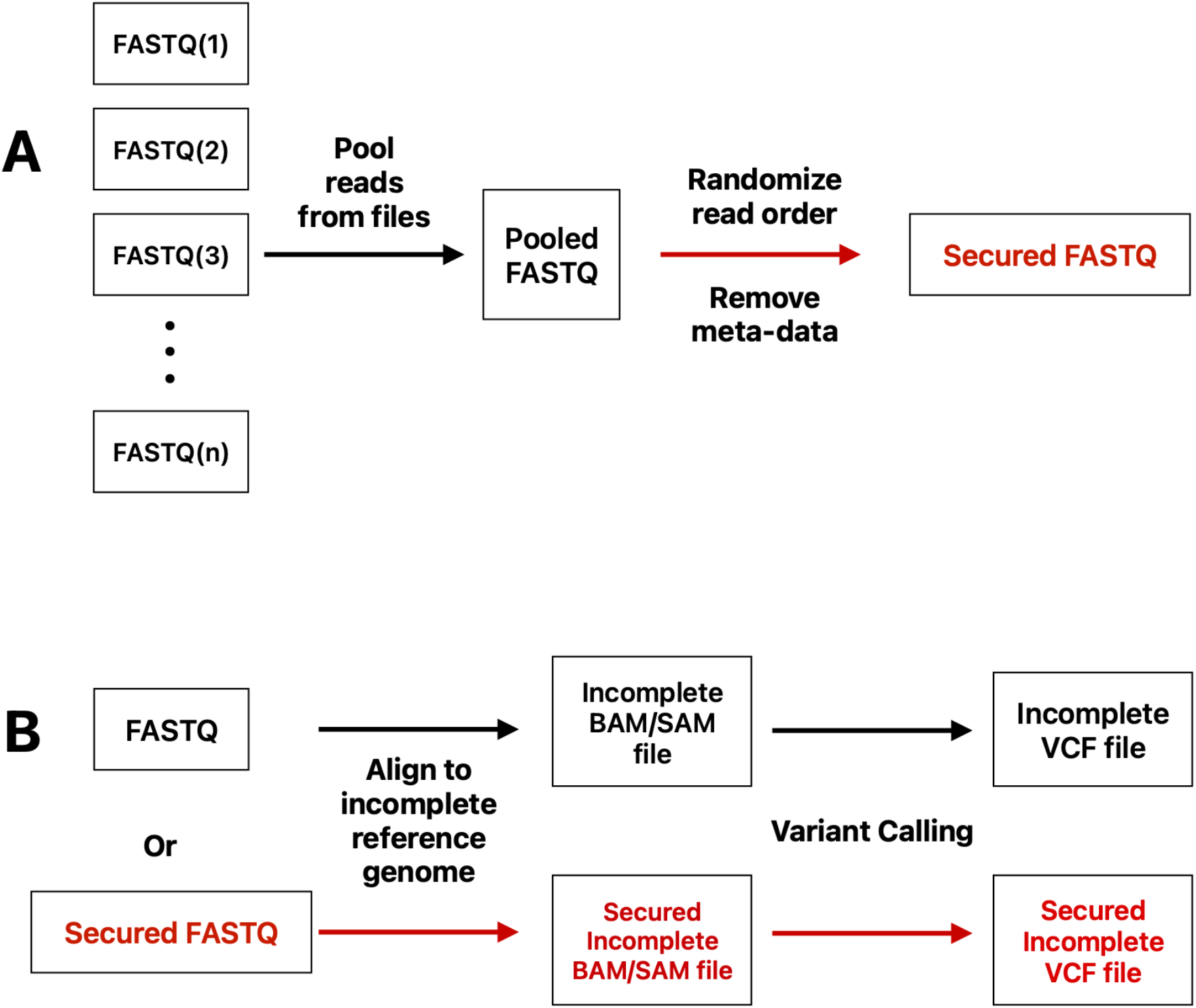
Workflow for methods used to obfuscate genomic data. **A. Method for pooling reads to generate a secure FASTQ file.** FASTQ files were pooled and their order randomized. **B. Method for removing genomic region(s) to limit information within a set of (pooled) samples.** Samples, or a pool of samples from (A), are aligned to an incomplete reference genome to produce a file that lacks complete genomic information.

The resulting alignment file (e.g. in SAM format) can then be processed into a variant call file (VCF) or can be shared directly. The aligned file or the VCF will be insufficient to recreate the full genome of the sample, as sensitive sequence regions that were removed are not included in the alignment of assembly files.

Our second proposed method is more sophisticated, and can occur prior to the application of the first proposed method (Figure 1A). Here, we pool DNA sequence reads from multiple samples of the same organism (e.g. in FASTQ format) and the metadata from each read is stripped. The order of the reads in the pooled data is then randomized. The resulting file remains in FASTQ format and contains all of the reads from the data of the original samples but paired-end read information is lost. The files with pooled reads can be directly processed into VCF files by mapping reads to a reference genome and calling variants. For additional security, the pooled data can also be processed with the first proposed method (aligning to an incomplete reference genome).

To examine the read pooling method, we applied it to FASTQ data from SARS-CoV-2 and from *Bacillus anthracis*. For SARS-CoV-2, we generated 7 independent datasets in FASTQ format, consisting of a unique mixture of 2, 5, 10, 50, 100, and 500 samples obtained from CDC (see supplementary table 1). For *B. anthracis* we generated 4 distinct datasets, consisting of 2, 5, 10 and 50 isolates, randomly subselected from NCBI SRA. (see supplementary table 2) with search criteria “Organism = Bacillus anthracis” and platform = Illumina” and “layout = paired”.

The metadata from each sample, including the depth of coverage and strain variants/lineages, were utilized as the ground truth for the composition/abundance of each pooled dataset. For SARS-CoV-2, the EDGE COVID-19, build 20230131 [22], was used for variant calling and lineage assignment was done with Pangolin (v4.3.1)[23]. For *B. anthracis*, the EDGE, build 20231115 [24] was used for read-mapping to the reference genome *Bacillus anthracis* str. ‘Ames Ancestor’ (AE017334.2) and for variant calling.

Freyja v1.4.8 [25] was used for deconvoluting the variants/lineages in the datasets from the VCF files from the pooled SARS-CoV-2 data. This program was designed to deconvolute genomes for ‘mixed’ samples, with the ability to predict the SARS-CoV-2 lineages present within the mixed samples. We used Freyja to predict the proportion of SARS-CoV-2 variants and lineages in the datasets, for both pooled data and for individual samples.

To examine the pooled *B. anthracis* data, we generated variant frequency heatmaps using the Plotly dash-bio.Clustergram package [26]. The Euclidean pairwise distance [27] and complete linkage clustering algorithms [28] were employed to the variant calling result based on variant locations. By applying these algorithms to the variant calling results, we can identify groups of variants with similar frequency patterns across different genomic locations. This clustering approach helps to identify, visualize, and interpret the relationships between variants in the *B. anthracis* data, potentially revealing insights into genetic similarities or differences among the different treatments.

## Results

By examining pooled data for larger and larger pools of samples (FASTQ reads), we tested the ability of our pooling method to obfuscate genomic data using traditional bioinformatics methods. We estimated the composition of the pooled data using these methods and compared the results to the true composition based on the samples used to generate those pools. For the SARS-CoV-2 samples, the EDGE COVID-19 workflow was used to provide majority-rule based variant calls [22]. Consequently, regardless of whether the SARS-CoV-2 reads were pooled or whether samples were analyzed separately, the same single nucleotide polymorphism variants (SNVs) and insertion/deletion (INDELs) were found. Freyja, another tool designed for SARS-CoV-2 [25], provides a population level analysis of the pooled reads. Freyja was successfully able to estimate the proportion of variants present within the pooled data (Figure 2). Note that the samples used to generate the pool of 5 datasets did not contain any Delta variant, being entirely composed of the Omicron variant, so a comparison of variant calling was not possible with this group.

**Figure 2.**
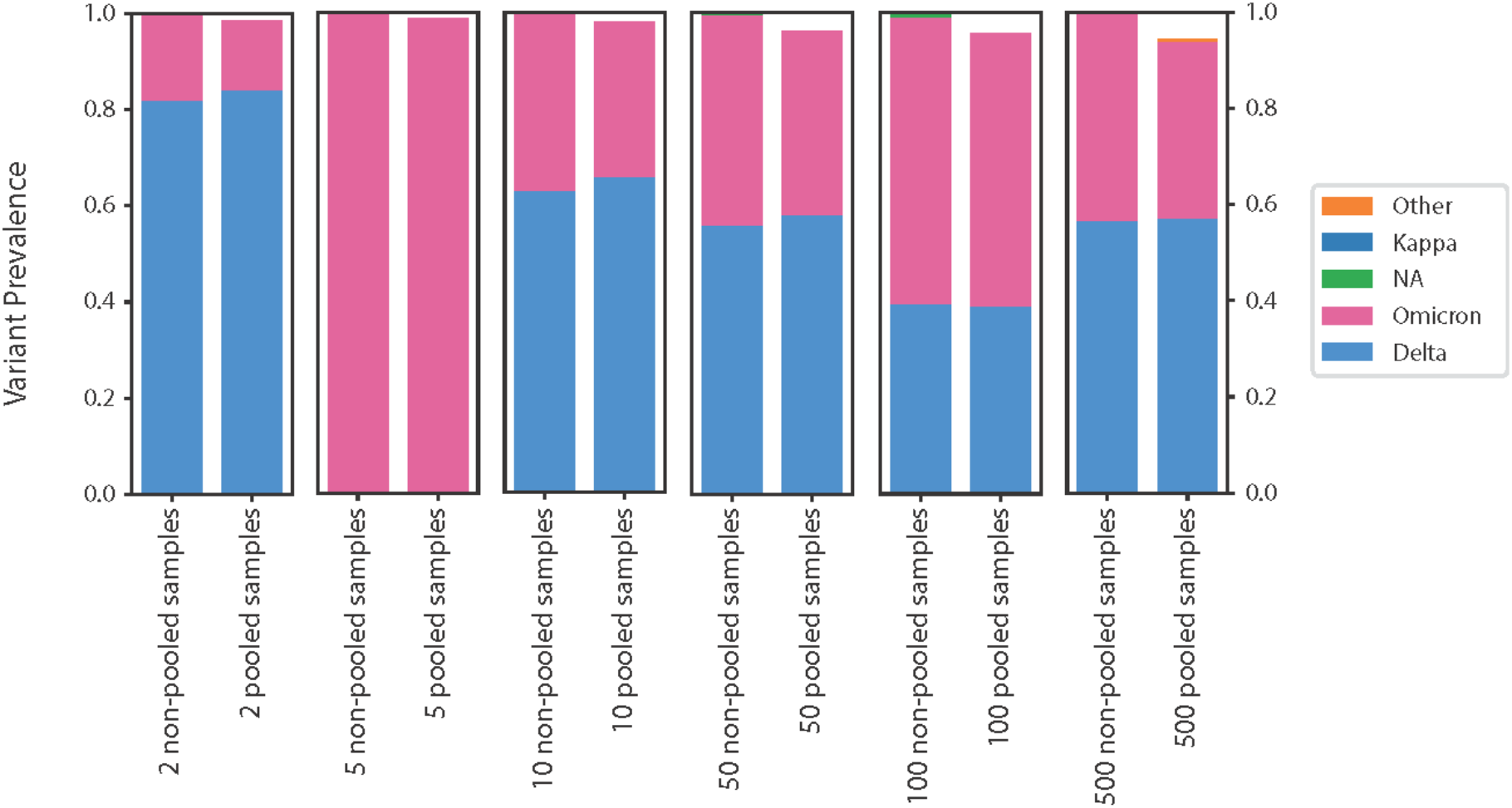
Prediction of variants present for each sample and for the pooled datasets. For each group examined, Freyja was used to predict variants present within each individual sample (left stacked histogram in each subplot), and the proportion of each variant in the pooled sample represents the ratio of reads from that sample used in the pooled dataset. The right stacked histogram for each subplot represents the variants present within the pooled datasets as predicted by Freyja.

While variants were accurately detected using the lineages identified by Freyja were not always correct (Figure 3). The prediction of lineages in Freyja produced primarily false positives, even when only two samples were pooled. Freyja’s ability to predict the presence of lineages in pooled data had diminishing accuracy as more samples were pooled together. Notably, Freyja tended to overestimate the number of lineages in the pooled data, and this overestimate increased as more samples were pooled together.

**Figure 3.**
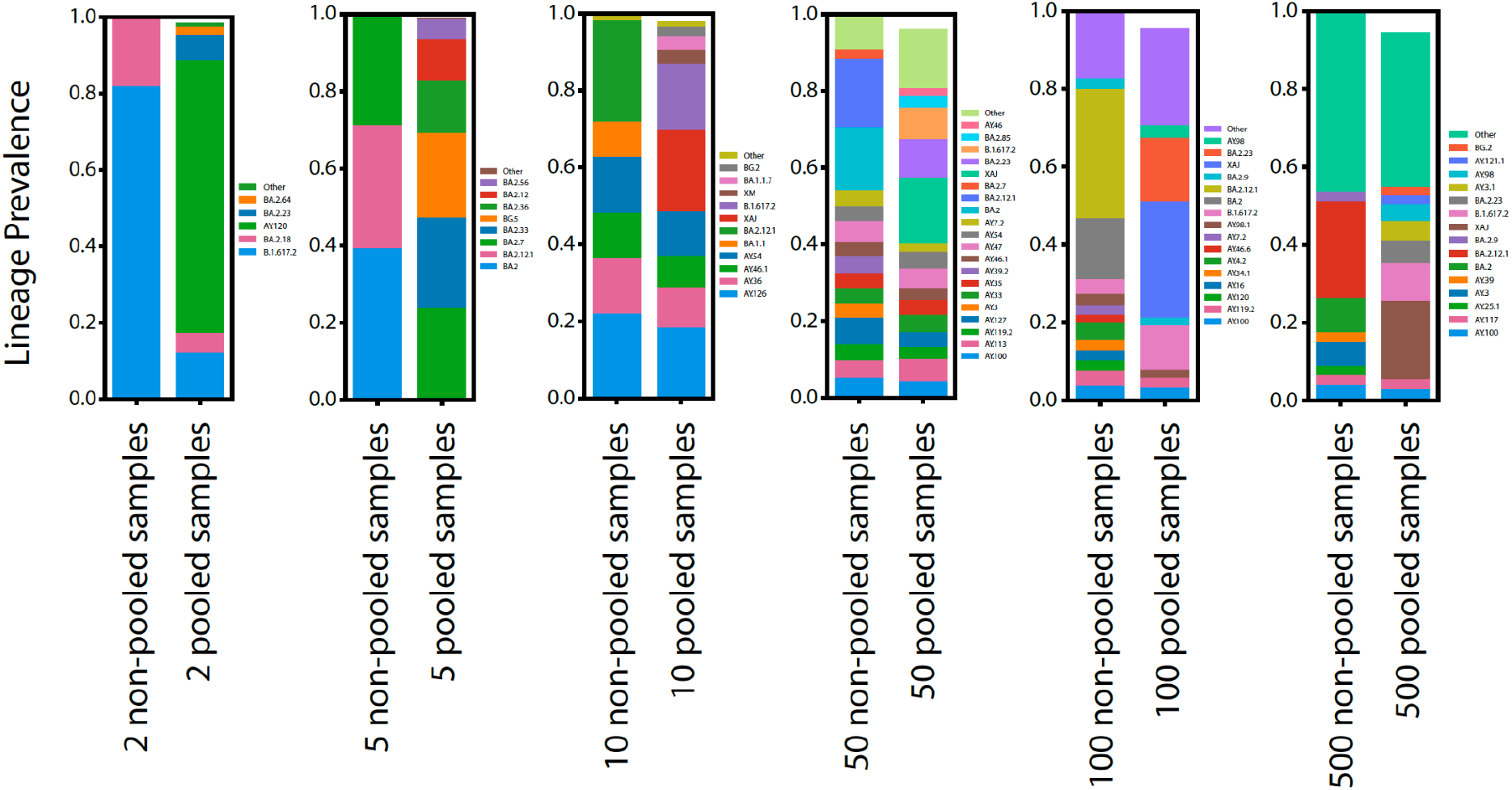
Prediction of lineages present within samples and within the pooled datasets. For each group examined, Freyja was used to predict lineages present within each individual sample (left stacked histogram in each subplot), and the proportion of each lineage in the pooled sample represents the ratio of reads from that sample used in the pooled dataset. The right stacked histogram for each subplot represents the lineages present within the pooled datasets as predicted by Freyja.

While the community is familiar with delineating variants in viral outbreaks, the definition of a lineage within bacterial species is not as straightforward. Due to the lack of sublineage definitions, and also, since there exists no similar program to Freyja for bacteria, we took a different approach to demonstrate that our method provides obfuscation of bacterial genomes.

For *B. anthracis* we generated 4 distinct datasets, consisting of 2, 5, 10 and 50 isolates, for a total 67 SRA samples. When the 67 individual samples of *B. anthracis* were individually mapped to the reference genome, an average genome coverage ranging from 97.04% to 99.06% was achieved. The variant calling analysis revealed a SNV count ranging from 8 to 2,653. To assess the pooled *B. anthracis* data, we compared the SNV positions in the pools with those from individual samples. Figure 4A presents a Venn diagram illustrating the SNVs found in the pooled 2-samples relative to those found in the individual samples alone (SRR5811050 with a mean depth 54x and SRR13950249 with a mean depth 203x). The pooled samples share 1,115 SNVs with the individual samples. However, the pooling of two samples introduced three false positive SNVs that were not present in the individual sample analyses. These false positives occurred because, although the alternative base ratios in the individual samples were below the detection threshold at these positions, their combined ratios in the pooled sample exceeded the threshold and were detected by the variant calling algorithm. Conversely, each individual sample contains 1,307 SNVs that were not detected in the pooled data, representing a high false negative (FN) rate in the pooled analysis. This high rate is likely due to two factors: (1) pooling increased the depth coverage of non-variant regions, diluting the signal of true variants (1,196 FN in SRR13950249 and 53 FN in SRR5811050), and (2) the pooling method removed paired-end information, preventing the mapping algorithm from rescuing reads that did not perfectly align due to SNVs (58 FN shared by two individual samples). Figure 4B depicts a cluster analysis heatmap of the pooled 2-samples data, along with data from the two individual samples. The vertical dendrogram on the left of Figure 4B illustrates the hierarchical clustering of the samples, revealing patterns of similarity. The heatmap itself shows variations in alternate base ratios across different SNV positions, with color gradients indicating the frequency of these variants. As seen in Figure 4B, although the SNV positions in the pooled samples overlap with those in the individual samples, there are differences in the alternative base ratios for other SNV positions.Notably, the top cluster highlights a “dilution” effect for false negative SNVs, where pooling reduces variant signals below detection thresholds. This result demonstrates that combining reads from different samples can obscure variant information, leading to both false positives and false negatives.

**Figure 4.**
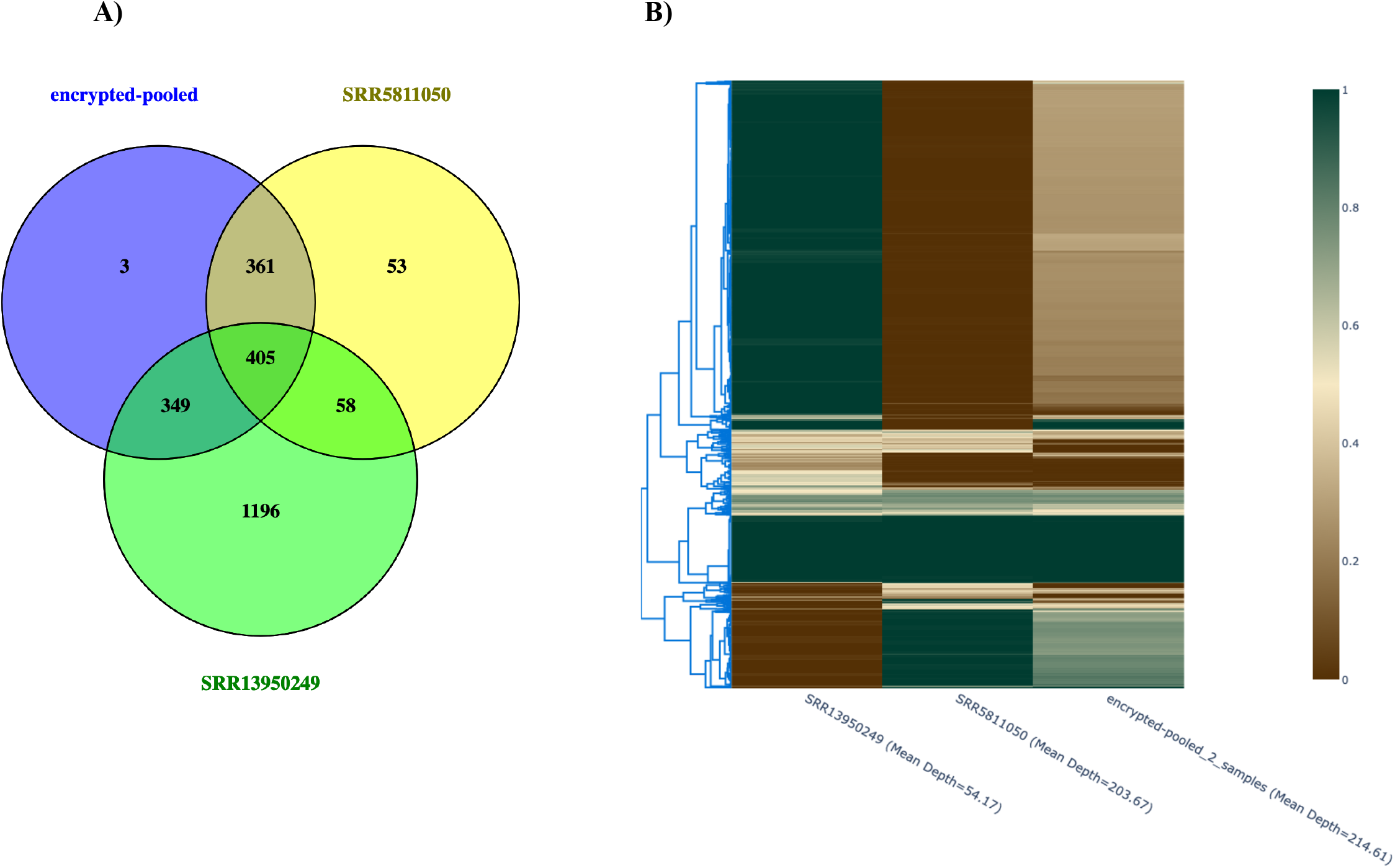
Pooled 2-samples SNVs and individual sample SNVs. A. Venn diagram. (https://bioinfogp.cnb.csic.es/tools/venny/.) The variants present within two individual samples (SRR5811050 and SRR13950249) were compared to determine which variants overlap between the different samples and also between the variants called in the pooled samples. **B. Heatmap.** Y-axis represents SNV position clustering. Heatmap color represents the SNVs ratio which is defined as the alternate base versus the reference base ratio.

## Discussion

The data generated by our software has a similar format to a real-life scenario in which the read data from individual samples is not clear. Samples can contaminate each other, multiple variants of a disease can co-infect a patient, or a new, recombinant lineage can arise. The program Freyja is designed to disentangle these potential scenarios for SARS-CoV-2, making it an ideal approach by which we can test the limits of our method. Freyja was unable to accurately predict which SARS-CoV-2 lineages were present within our pooled data, indicating that our method successfully obfuscates lineage information (Figure 3).

Consequently, our pooling method can prevent the reconstruction of complete SARS-CoV-2 genomes, even when only a few genomes are pooled. Freyja’s ability to accurately estimate the fraction of variants in our pooled data, on the other hand, indicates that our method does not conceal lineage-specific information (Figure 2).

When constructing genomes, many factors can influence variant calling. We expected that the differences between the bacterial and viral genomes examined here would influence how well our pooling method would obfuscate the genomes. Bacterial genomes are larger and more structurally complex than viral genomes and contain more variation, and thus the reconstruction of their genomes can be more difficult. Furthermore, delineation of bacteria lineages can be complicated by frequent horizontal gene transfer from distant species and large-scale genome rearrangements [29,30]. Therefore, we expected *a priori* that reconstructing *B. anthracis* genomes from the pooled sequence data would be a bigger challenge than for SARS-CoV-2. Without a custom tool like Freyja to disentangle genomes, our analysis instead focused on a comparison of variant calling between pooled data and individual samples.

For data pooled from two *B. anthracis* samples, the SNPs called in the pool did not always match the SNPs independently called from the two original samples (Figure 4A, 4B). These results demonstrate that pooling data obfuscates variant calling. Accurate variant calling is needed to fully reconstruct genomes or to predict the presence of specific bacterial lineages. This effect becomes more pronounced as additional samples are pooled (see supplementary figures), making it increasingly difficult to accurately call variants and fully reconstruct genomes.

Overall, these results demonstrate that by pooling reads from the same species, the samples’ genomes become obfuscated. Not all information is concealed, however. Much of the genomic information remains: the species being sequenced, the common variants that are present in the pool, and general information about gene sequences. We also found that pooling more samples always increases genome obfuscation, and larger genomes with more variants are more difficult to accurately reconstruct. However, the limits of this method have not been tested here. Pooling genomes of distantly related organisms will facilitate the attribution of a specific read to a specific lineage, limiting obfuscation of the samples of distant relation to the rest of the pool. Furthermore, if two samples are nearly identical in their genome sequence, then pooling their sequence data will do little to change the overall composition of the pool.

Additional work is required to determine exactly how sequence diversity, genome size, and the number of pooled samples affect how information is concealed with our method. To address this uncertainty, aligning reads to an incomplete reference genome (Figure 1A) can always be used to ensure that full reconstruction of a genome is not possible.

Many other types of genetic data are widely shared. For example, human genomic data can be shared to further research on human genomics or to develop new pharmaceutical drugs [31]. The sharing of human genomic data is complicated by issues of identifiability of the data [32] and the consequent violations of privacy [33,34,35]. Human genetic data also has potential security issues that result from the fact that the people from whom the data is generated will be alive for decades and future technological advancements in biotechnology and AI may facilitate the misuse of this data [36]. Like with pathogen data, concerns about the insecure sharing of human genomic data may ultimately lead to limitations on how data is shared. The application of our data security techniques to human data represent a potential future direction of this work.

## Conclusion

The genomes analyzed here are of importance to biosecurity, and this method can have widespread applications to genomic data of other pathogens. This method can also be applied to larger genomes, such as human, to obfuscate sensitive data for privacy and security. Pooling genomic data, or equally, pooling genomic material, results in a mixture that prevents full reconstruction of a genome. This method, potentially combined with methods that remove some regions of the genome, can nevertheless allow useful genomic data to be shared. The computational methods used here also have the potential to guide and optimize methods in molecular cryptography. The effectiveness of different pooling methods can be examined with this software, and the results can be used to design molecular cryptography.

By sharing raw data in a way that prevents misuse, analyses can be done to ensure an accurate interpretation of the data. The use of genetic data, in pathogen surveillance and elsewhere, can be essential for biosecurity, for the development of the bioeconomy, and the advancement of medicine. This data sharing can be stifled by the threat of misuse. The availability of raw data can help ensure the accuracy, authenticity and interpretation of any genomic analyses. Even when raw data is not to be shared, securing information within raw data itself provides security for the data at rest, and ensures that any information extracted from the data remains equally secure.

## Data Availability

All data produced are available online at the NCBI SRA repository.

## List of abbreviations

DNA: Deoxyribonucleic acid
SARS-CoV-2: Severe acute respiratory syndrome coronavirus 2
CRISPR: Clustered regularly interspaced short palindromic repeats
VCF: Variant Call File
FN: False Negative

## Declarations

### Ethics approval and consent to participate - Not applicable Consent for publication - Not applicable Availability of data and materials

The datasets analyzed during the current study are available in the NCBI SRA repository and their accession numbers are listed in supplementary table 1 and table 2.

### Authors Contributions

SS, BH and PC designed the research plan. CCL and PL managed the data and conducted the analyses. All authors contributed to the writing of, read and approved the final manuscript.

## Acknowledgements

**Not applicable**

