## Supplementary material for "Methods for safely sharing dual-use genetic data": SupplentaryFigures

**Supplementary Figure 1. Pooled 5-samples SNVs and individual sample SNVs**

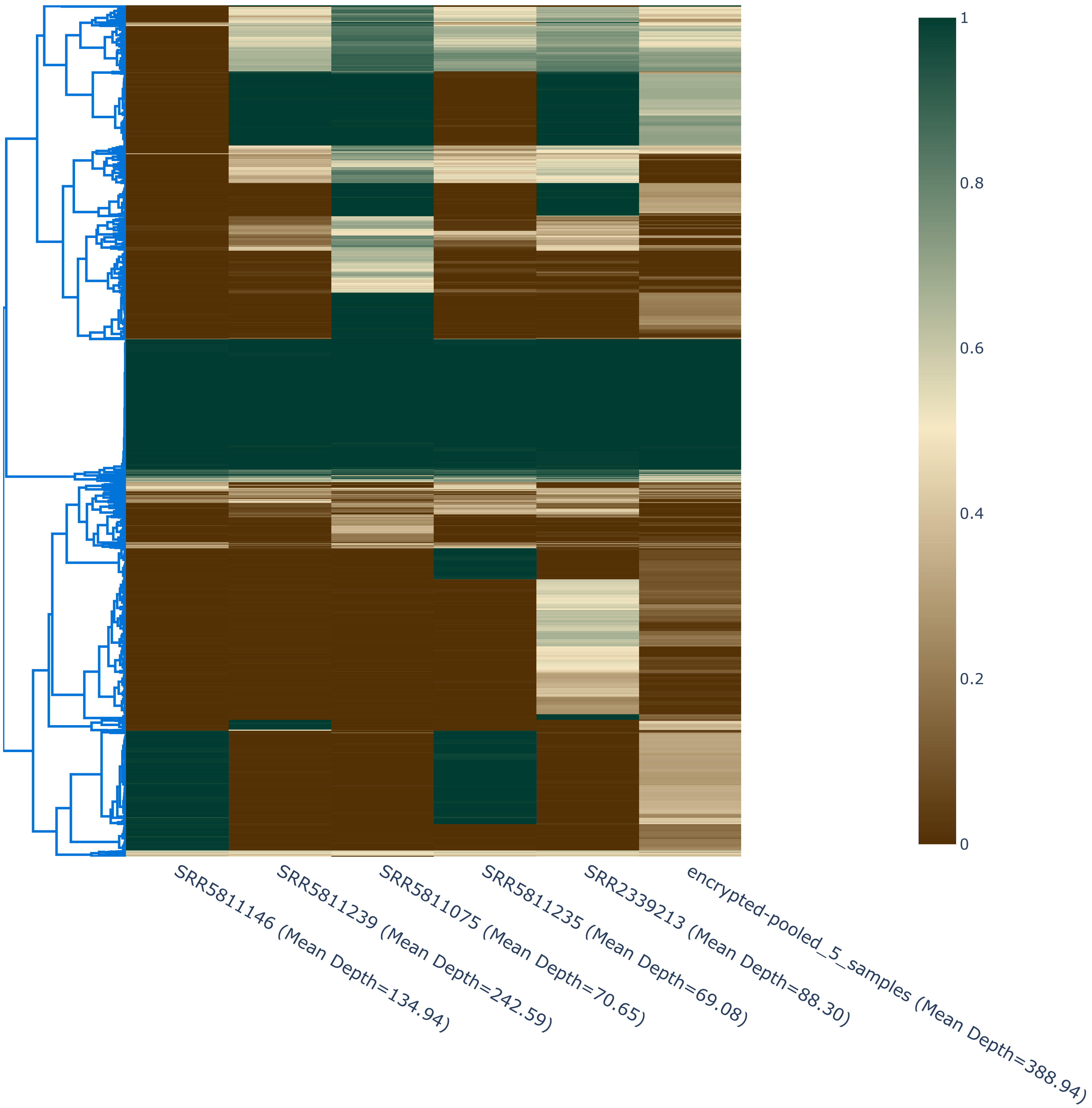

**Supplementary Figure 2. Pooled 10-samples SNVs and individual sample SNVs**

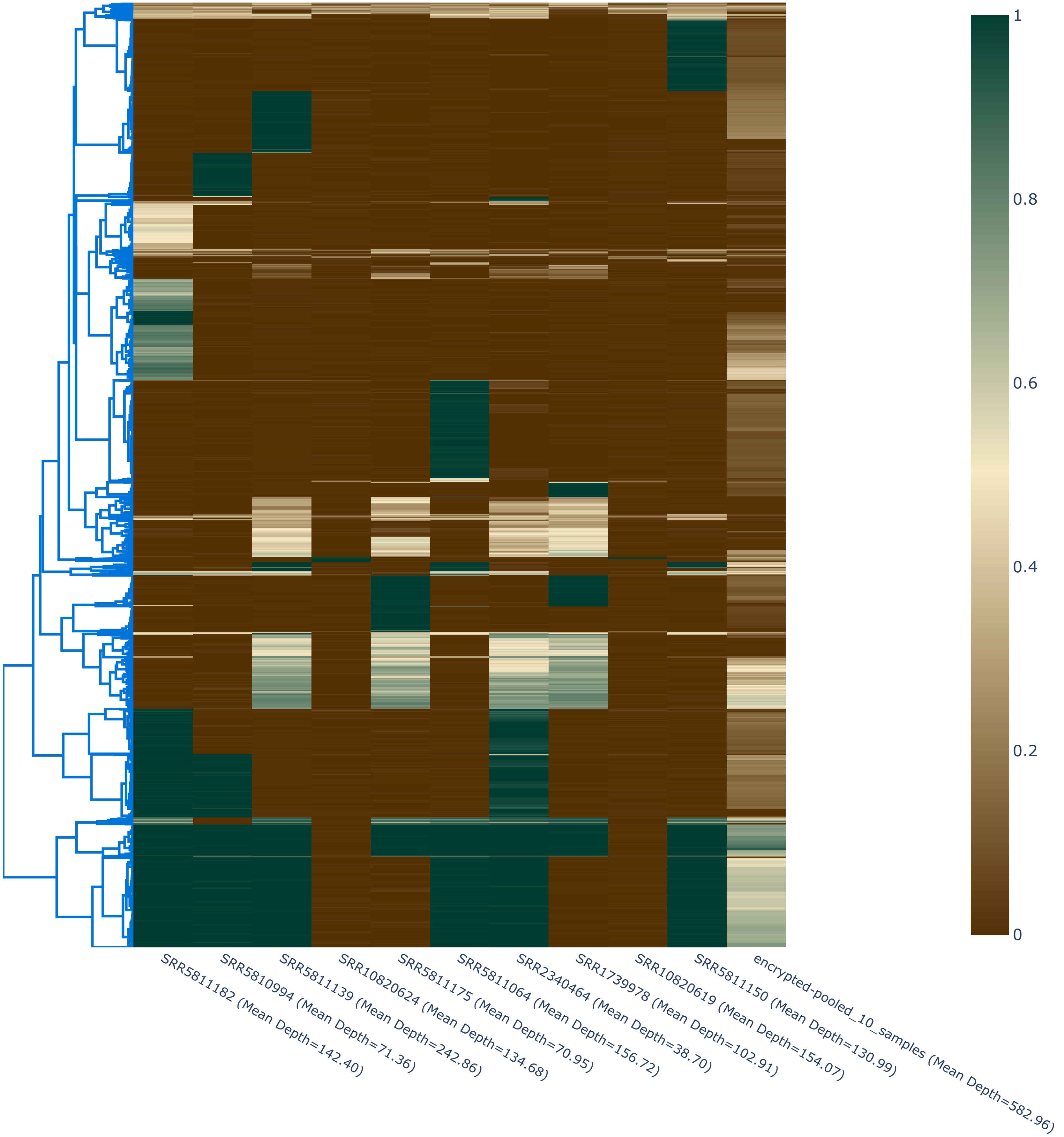

**Supplementary Figure 3. Pooled 50-samples SNVs and individual sample SNVs**

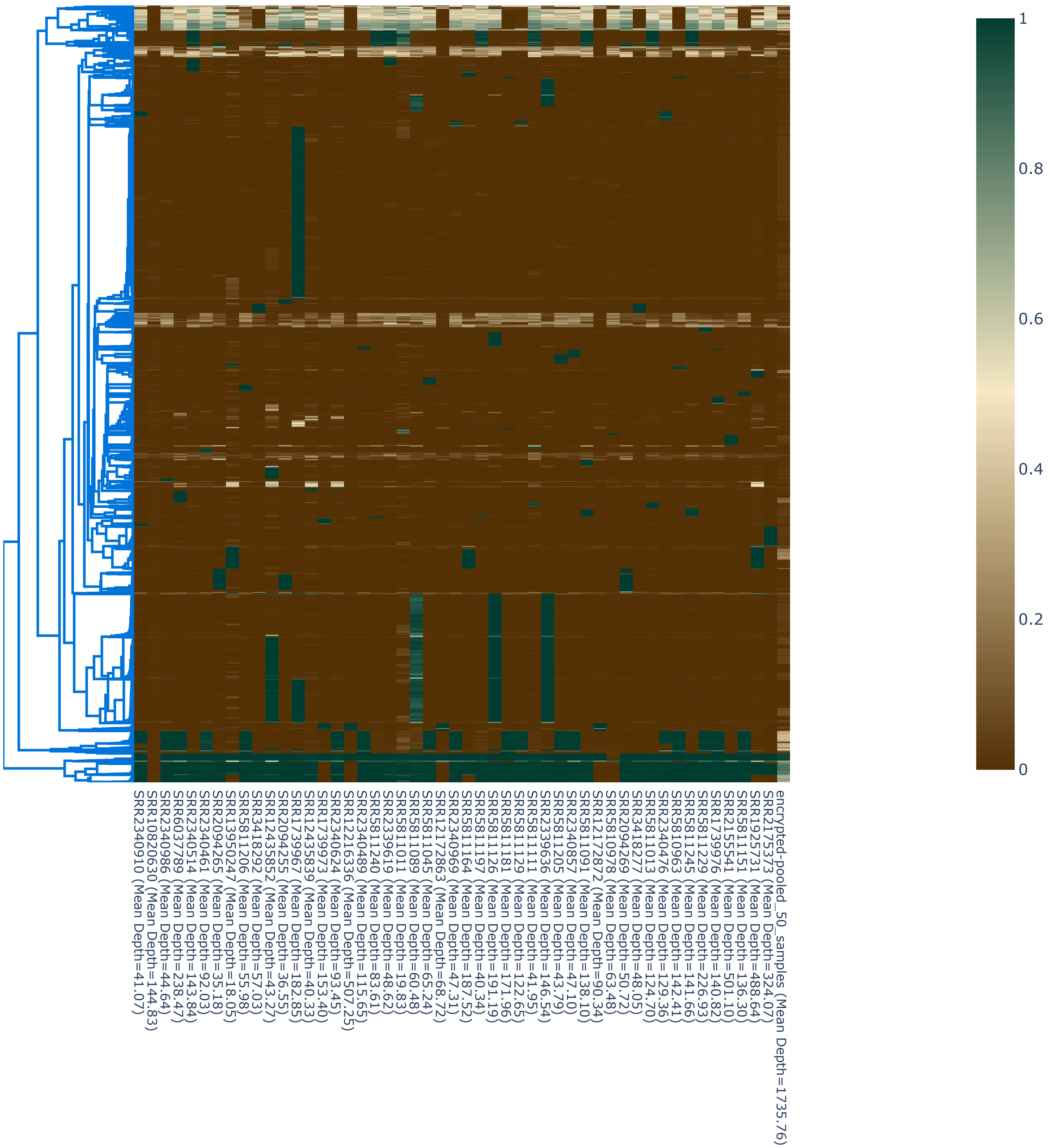
